# Development and preliminary testing of an online tool to assess social inclusion and support care planning in mental health supported accommodation

**DOI:** 10.1101/2023.01.03.23284140

**Authors:** Sharon Eager, Helen Killaspy, C Joanna, Gillian Mezey, Peter McPherson, Megan Downey, Georgina Thompson, Brynmor Lloyd-Evans

## Abstract

**Background:** Individuals with severe mental illness living in supported accommodation are often socially excluded. Enabling social inclusion is an important aspect of recovery-based practice, and improves quality of life. The Social Inclusion Questionnaire User Experience (SInQUE) is a measure of social inclusion that has been validated for use with people who have mental health problems. Previous research has suggested that the SInQUE could also help to support care planning focused on enabling social inclusion in routine mental health practice.

**Objectives:** To develop an online version of the SInQUE for use in mental health supported accommodation services, and examine its acceptability and perceived usefulness as a tool to support care planning with service users.

**Methods:** i) A lab-testing stage to assess the acceptability of the SInQUE tool through ‘think-aloud’ testing with six supported accommodation staff; ii) A field-testing stage to assess the acceptability, utility, and use of the SInQUE tool over a 5-month period. An implementation strategy was employed in one London borough to encourage the use of the SInQUE. Qualitative interviews with 12 service users and 12 staff who used the tool were conducted and analysed using thematic analysis. Use of the SInQUE was compared with two other local authority areas, one urban and one rural, where the tool was made available for use but no implementation strategy was employed.

**Results:** In total, 17 staff used the SInQUE with 28 different service users during the implementation period (about 10% of all service users living in supported accommodation in the study area). Staff and service users we interviewed felt that the SInQUE was collaborative, comprehensive, and user-friendly. Staff deemed the tool relevant to their role. Although some staff were concerned that particular questions might be too personal, service users did not echo this view. Participants generally felt that the SInQUE could help to identify individuals’ priorities regarding different aspects of social inclusion through prompting in-depth conversations and tailoring specific support to address areas where service users would like to be more included. Some interviewees also suggested that the tool could highlight areas of unmet or unmeetable need across the borough, that could feed into service planning. The SInQUE was not used in the comparison areas that had no implementation strategy.

**Conclusions:** The online SInQUE is an acceptable and potentially useful tool, that can be recommended to assess and support care planning to enable social inclusion of people living in mental health supported accommodation services. Despite this, take-up rates were modest during the study period. A concerted implementation strategy is key to embedding its use in usual care, including proactive endorsement by senior leaders and service managers.

## Introduction

Social inclusion refers to an individual’s ability to participate in important societal activities and their sense of community belonging [1,2]. Someone may feel socially excluded if they do not have opportunity for societal involvement and integration, often due to external factors that are beyond their control [3]. Social exclusion is a multifaceted continuum [2], typically signified by poverty, unemployment, inequality, and/or poor health [4]

People with serious mental illness are thought to be among the most socially excluded groups in society [5]. Individuals with these kinds of mental health problems often have smaller and less satisfying social networks [6], lower household income [7], lower levels of employment [8] [9], and experience more criminal and violent victimization [10,11] than those in the general population. Social exclusion can be conceptualized as both a cause and a consequence of mental ill-health [12]. Furthermore, greater social inclusion is associated with better quality of life and lower levels of loneliness among those with severe mental illness, suggesting that social exclusion is an important area for mental health practitioners to try to address [13,14].

Mental health supported accommodation services provide care and support to individuals with particularly severe and complex mental health problems, as a way of supporting recovery in the community [15]. It is estimated that there are around 100,000 people living in mental health supported accommodation in England. Services are typically staffed by support workers, with additional, specialist clinical input provided from National Health Service (NHS) community mental health teams [16]. In England, three main types of supported accommodation are provided: i) residential care homes for those with the highest needs that comprise 24-hour staffed communal facilities where placements are not time limited, with meals, supervision of medication, cleaning, and activities provided for service users; ii) supported housing services that provide shared or individual, self-contained, time-limited tenancies with staff based on-site up to 24 hours a day to assist service users in gaining skills to move on to less supported accommodation; iii) floating outreach services that provide visiting support of a few hours per week to people living in permanent, self-contained, individual tenancies, with the aim of reducing support over time to zero [16].

Service users living in supported accommodation are often socially isolated, with low levels of employment and little involvement in civil and political processes [17]. Many report feeling lonely and isolated, and experience a high level of stigma that causes them to become more socially isolated [18]. There is evidence that users of mental health supported accommodation services report a variety of unmet needs, such as accessing employment opportunities and forming intimate relationships [19,20]. However, relatively little research has been conducted to inform the precise needs of service users living in supported accommodation [21], and a greater focus on this group is needed to identify and implement interventions that are likely to be the most useful for them [22].

Supporting service users to work towards desired goals and community engagement is highly congruent with recovery-based practice in mental health. Recovery-based practice recognises and builds on service users’ strengths and promotes empowerment through collaboration between them and staff to identify and work towards specific goals [23]. Many of the goals that are identified are markers of social inclusion, such as employment, social network development, and community activities [24]. Qualitative evidence from a large national research programme suggests that staff working in mental health supported accommodation services operate with a considerable degree of recovery orientation [24,25] and the more recovery-orientated these services are, the more likely people are to move on successfully to more independent settings [26].

People living in mental health supported accommodation have expressed a strong preference for individually tailored services that offer choice and promote autonomy, consistent with a recovery-based approach [18]. Patient-reported outcome measures have been recommended to inform this individualised approach, by directly capturing service users’ perspectives on constructs such as goal attainment, quality of life, and social inclusion [27]. Such measures enable service users to make informed decisions about their own support and care planning, in line with WHO recommendations for recovery-based practice in community care provision [28]. Online resources delivered across web-based platforms have been established as accessible, acceptable, and effective for use among participants with severe mental illness, particularly those which offer guided support [29,30]. A tailor-made online assessment tool, the Quality Indicator for Rehabilitative Care - Supported Accommodation (QuIRC-SA) has also been successfully used by managers of supported accommodation services, suggesting that these settings have the required resources and expertise to implement online measures [31].

The SInQUE was developed as a measure of social inclusion for individuals with severe mental illness [32]. The measure is validated across a range of mental health populations, has established reliability, is considered acceptable to service users, and has been proposed as potentially cross-culturally suitable [32–34]. Thus far, the SInQUE has been used solely in offline, research contexts. However, stakeholder feedback from a prior study testing the SInQUE indicated that the measure may be useful in clinical practice to assess social inclusion, facilitate important conversations with service users, and guide care and support planning [34]. Furthermore, a consistent research recommendation from the developers of the SInQUE tool has been to investigate whether the measure has utility as a care planning tool, to promote social inclusion in routine mental health practice [13,34].

The present study aimed to develop an online version of the SInQUE for use in mental health supported accommodation services. We sought to examine the acceptability and perceived usefulness of this tool among supported accommodation staff and service users, as a means to assess their needs for greater social inclusion and promote care planning.

### Aims

The SUSHI study aims were:

1. To develop and refine an online version of the SInQUE social inclusion assessment tool, tailored for use in mental health supported accommodation settings.
2. To investigate the acceptability and perceived utility of the online SInQUE tool among supported accommodation staff and service users.
3. To determine the extent of take-up of the tool in supported accommodation settings, with and without a locally developed implementation strategy to support its use.
4. Informed by study findings, to develop a programme theory and logic model for the online SInQUE; specifying its anticipated outcomes, the mechanisms through which they may be achieved, and contextual factors impacting use and experience of the SInQUE.

## Methods

### Study design

This study comprised two stages, conducted in one inner London borough:

1. A lab-testing stage to assess initial acceptability of the tool and develop it through ‘think-aloud’ testing and semi-structured interviews with supported accommodation staff.
2. A field-testing stage to assess wider acceptability, feasibility, and usage of the tool. Semi-structured interviews were conducted with staff and service users who had used the online SInQUE during this stage.

The five-month field-testing stage was supported by a local implementation strategy, developed in collaboration with local service leads, to support use of the online SInQUE by supported accommodation staff in the participating London borough. The online SInQUE was also made available to supported accommodation services by local service leads in two other areas, without any accompanying implementation strategy.

### Description of the SInQUE tool

The online version of the SInQUE [35] can be used to assess social inclusion and inform support and care planning for people with mental health problems. It is designed to be used by staff as part of routine care planning, to be completed collaboratively with service users. It is compatible for use on a computer, tablet, or mobile device. Staff are required to register an account on the SInQUE site using their work email address and details of their organisation, and can then use the tool for free. No personal data identifying service users are logged or stored on the SInQUE platform. The online SInQUE generates a unique reference number for each new service user, which is retained by the staff member completing the assessment for future reference and to link any repeat assessments.

The online tool developed for lab-testing in our study included the 46-item version of the validated SInQUE social inclusion questionnaire, which was refined following stakeholder feedback at the end of the previous measure development study [34]. The questionnaire yields a total score of 0-75, with a higher score indicating greater social inclusion. Questions and sub-scale scores are grouped into nine different areas of social inclusion: leisure, social relationships, religious and cultural activities, education and employment, transport, health, crime victimisation, home life and housing, and civic duties. These areas cover the five social inclusion domains of the validated SInQUE (social integration, productivity, consumption, access to services, and political engagement), but the nine named areas listed above were considered more immediately understandable for use in practice.

Using the service user’s responses, the online SInQUE generates a list of areas where the person has said they would like to be more socially included. It then offers a prompt for the service user and staff member to collaboratively select up to three priority areas that they would like to integrate into the person’s support plan. Once complete, a summary report is generated. If the assessment is repeated with the same service user in the future, this report also displays changes in their social inclusion over time. The tool can generate management-level summary reports for each organisation that is registered with it, and commissioner-level summary reports of services using the tool across a whole area (such as a London borough). Please see the supplementary material (Appendix A) for a full description of the SInQUE using the Template for Intervention Description and Replication (TIDieR) checklist [36].

### Setting

This study took place in mental health supported accommodation services across one inner London borough. There are 21 such services in the borough run by six different voluntary sector organisations. They offer varying degrees of support to over 270 service users who are also supported by local NHS secondary mental health services. In the borough, there are approximately 24 service users living in residential care, 159 in supported housing, and 89 who receive floating outreach support. Supported housing services offer 24-hour support to 119 people and ‘9 to 5’ support to 40 individuals.

### Lab-testing stage: recruitment, data collection and analysis

Six supported accommodation staff members were recruited to provide initial impressions of the online SInQUE tool. We discussed the study with service managers working in three different services, and asked them to nominate two staff members each from their service who were interested in taking part. Participants were purposively sampled to include staff working in floating outreach support, 24-hour supported housing, and residential care.

Data were collected in January and February 2022. All six think-aloud interviews with staff were conducted and recorded via Microsoft Teams. The researcher first discussed the information sheet with each participant and gave them the opportunity to ask questions. Following this, participant consent was verbally collected and audio-recorded separately to the main part of the interview. Participants were then asked to fill out a short, online form, providing their demographics, before beginning the interview.

We conducted ‘think-aloud’ testing of the online tool with staff, using a semi-structured topic guide developed by the study team (Appendix B). Following a process used previously in developing online tools [37], participants were asked to complete set tasks using the online SInQUE tool while providing a continuous commentary on their thoughts. They were asked to open the SInQUE website, register an account, and complete an assessment as they would do with a service user. At all stages they were prompted to share their thoughts as they navigated the website and offer their initial impressions on how easy it was to understand and use, and its potential suitability for their work. Once participants had completed the questionnaire, the researcher asked some broader questions about their experience using the tool and any ways it could be improved.

Identified problems and suggestions for improvements to the online tool were collated by the researcher following the interviews. They were then reviewed by the study team and decisions about refinements to the online tool were agreed and the tool was revised accordingly.

### Field-testing stage: recruitment, data collection and analysis

#### 1. The Local Implementation Strategy

The revised version of the tool was made available for use in mental health supported accommodation services across the participating inner London borough. We iteratively developed and implemented a strategy to encourage and support its use by supported accommodation staff in the borough over a five-month period beginning 11^th^ of May 2022. This implementation strategy was informed by consultation with supported accommodation service managers, clinicians working in the Islington community mental health rehabilitation team, and from the individual interviews conducted with supported accommodation service users and staff.

#### 2. Interviews with field-testing participants

### Participants and recruitment

Individual interviews with service users (n=12) and staff (n=12) who tried out the SInQUE tool were conducted from late May to September 2022. This number was chosen to explore the views of staff and service users from a variety of supported accommodation types and service providers. Once a staff member informed the study researcher that they had tried the tool in practice, we asked them whether they would like to participate in an individual interview about their experience. We also invited staff to pass on information about the study to the service user(s) with whom they had used the tool, and to ask them whether they would like to participate in an interview about their experience.

We recruited these participants purposively from a range of supported accommodation types and provider organisations. One service user interview and one staff interview were conducted online via Microsoft Teams; all other interviews were carried out in person, according to participants’ preference. In-person interviews were conducted by the study researcher at the staffed supported accommodation sites, aside from one interview with a service user receiving floating outreach support, which was conducted at their home.

### Measures and procedures

The researcher first discussed the information sheet with participants and gave them the opportunity to ask questions about the study. For in-person interviews, informed consent was collected via paper consent form; for online interviews verbal consent was audio recorded. Participants were then asked to answer some brief demographic questions about themselves and their associated service. Following this, the researcher asked each participant questions about their experience using the SInQUE, if there were any ways it could be improved, the appropriateness of the online tool for use in their work, and what impacts, if any, they thought it might have on care provision and service users’ experience. The interview topic guides (one for staff participants and one for service user participants) were developed by the study team as semi-structured interviews: they are provided in the supplementary material (Appendix C). In-person interviews were recorded using a digital voice-recorder; online interviews were recorded on Microsoft Teams. Service user participants were offered a £20 shopping voucher to thank them for their time. Interview audio-recordings were transcribed by a professional transcription company, with which University College London (UCL) had a data sharing and privacy agreement. Interview transcripts were then checked by the study researcher for accuracy. Any potentially identifying text was anonymised. The resulting, cleaned transcripts were then securely stored on the UCL university system.

### Analysis

Analysis of the interviews comprised two stages. Firstly, the study researcher noted any problems or improvements to the online SInQUE suggested by participants. These were then reviewed by the study team and changes to the online SInQUE agreed, as in the previous lab-testing phase.

Secondly, transcripts were uploaded to NVivo (version 12) for qualitative analysis. As we aimed to develop a programme theory for the online SInQUE intervention, we initially coded data into a deductively derived framework which used an intervention-context-actor-mechanism-outcome (ICAMO) configuration, with each component of this ICAMO framework representing a primary theme [38]. Within each of these five primary themes, we then inductively derived subthemes from the data using thematic analysis. The initial coding was conducted by the lead author (SE) and was then reviewed and adjusted collaboratively by the study team. This included gaining lived-experience perspectives from a researcher with experience of mental health service use (JC) and clinical insight from a senior clinical academic working in the participating borough as a consultant rehabilitation psychiatrist, supporting service users who live in supported accommodation (HK). The team brought further perspectives from those with backgrounds in social work (BLE), clinical psychology (PM), forensic psychiatry (GM), and from the community rehabilitation team in the borough (MD).

#### 3. Data usage monitoring

Data on the uptake and use of the online SInQUE tool were collected from the online SInQUE informatics for the five-month field-testing period from 11^th^ May – 11^th^ October 2022.

At the start of this period, the study team also contacted local mental health service rehabilitation and housing leads in two other areas: another inner London borough and a rural county in the west of England. These service leads contacted local supported accommodation managers and invited them to use the online SInQUE in their service if they wished. The tool was made available to seven supported accommodation services in the London borough, and ten in the rural county. No further encouragement to use the tool or implementation support was provided. This allowed us to monitor take-up and use of the tool in two areas without an associated implementation plan and thus make inferences about the necessity and impact of the strategy we developed.

#### 4. Logic model development

The study team had developed a preliminary logic model for the online SInQUE in planning this study. We used the findings from the research activities described above to review and refine this logic model, and develop an updated theory about the potential outcomes for service users and organisations from using the online SInQUE, mechanisms through which these outcomes are achieved, and factors influencing the take-up, experience, and impact of the online tool. Factors related to: i) the intervention itself, ii) the characteristics and attitudes of staff and service users using the online tool, and iii) the broader organisational and societal context. This was summarised in a logic model in the form of an ‘ICAMO map’ [38], which was developed and refined iteratively through discussion with the study team.

### Ethical approvals

The initial lab-testing phase of this study (SUSHI Phase 1) was approved by the UCL Research Ethics Committee (REC) on 18/06/21 (REC reference 6711/002). The following field-testing phase (SUSHI Phase 2) was approved by the London Camden and Kings Cross NHS Research Ethics Committee on 04/11/2021 (REC reference 21/LO/0657).

## Results

### Participants

We recruited six supported accommodation staff members for the ‘think-aloud’ interviews during the lab-testing stage. We recruited twelve further staff members and twelve supported accommodation service users for the individual interviews as part of the field-testing stage. Participant characteristics for both stages are summarised in Table 1.

**Table 1.** Lab-testing and field-testing of the online SInQUE: characteristics of participants.

| Participant characteristics | Lab-testing: staff<br>(N=6) | Field-testing: staff<br>(N=12) | Field-testing:<br>service users<br>(N=12) |
| --- | --- | --- | --- |
| **Gender** |  |  |  |
| Male | 2 (33%) | 5 (42%) | 11 (92%) |
| Female | 4 (67%) | 6 (50%) | - |
| Non-binary | - | 1 (8%) | 1 (8%) |
| **Age** |  |  |  |
| 18 – 30 | - | 8 (67%) | 4 (33%) |
| 31 – 50 | 2 (33%) | 2 (17%) | 3 (25%) |
| 51+ | 4 (67%) | 1 (8%) | 4 (33%) |
| Prefer not to say | - | 1 (8%) | 1 (8%) |
| **Ethnicity** |  |  |  |
| White/White British | 2 (33%) | 8 (67%) | 1 (8%) |
| Black/Black British | 3 (50%) | 2 (17%) | 4 (33%) |
| Asian/Asian British | 1 (17%) | - | 1 (8%) |
| Mixed/multiple ethnic groups | - | 1 (8%) | 6 (50%) |
| Other ethnic background | - | 1 (8%) | - |
| **Sexual orientation** | N/A<sup>a</sup> | N/A<sup>a</sup> |  |
| Heterosexual/straight | - | - | 6 (50%) |
| Gay/lesbian | - | - | 1 (8%) |
| Bi/Bisexual | - | - | 2 (17%) |
| Prefer not to say | - | - | 3 (25%) |
| **Type of supported accommodation**<br>**lived/worked in** |  |  |  |
| Floating outreach support | 2 (33%) | 2 (17%) | 2 (17%) |
| 9 to 5 supported housing | 2 (33%) | 1 (8%) | 2 (17%) |
| 24-hour supported housing | 2 (33%) | 7 (58%) | 8 (67%) |
| Residential care | - | 2 (17%) | - |
| **Length of time worked/lived in**<br>**supported accommodation** |  |  |  |
| Less than 2 years | 1 (17%) | 7 (58%) | 5 (42%) |
| 2 – 5 years | 3 (50%) | 3 (25%) | 5 (42%) |
| 6 – 10 years | 1 (17%) | 2 (17%) | - |
| 10+ years | 1 (17%) | - | 1 (8%) |
| Prefer not to say | - | - | 1 (8%) |

Reported in the format N(approximate %), where N=number of participants.
<sup>a</sup>Staff were not asked about their sexual orientation

#### 1. Changes made to the SInQUE

Following phase 1 lab-testing and phase 2 field-testing, suggestions that participants made for how the tool could be improved were collated and reviewed by the team. Accordingly, adjustments were made to the online SInQUE after each stage, an overview of which can be found in Table 2.

**Table 2.** Changes made to the online SInQUE following phase 1 and phase 2 testing.

|  | Section of SInQUE affected | Explanation of the problem | Resolution | Justification for change |
| --- | --- | --- | --- | --- |
| **Registration and usage changes.** | The online SInQUE home page. | Some staff and service users suggested developing additional materials to help explain the SInQUE.<sup>a</sup> | Developed a guidance manual for service managers and commissioners with information on using the SInQUE. Also developed an informational leaflet and poster aimed at service users about the SInQUE. | Easier for managers, commissioners, staff, and service users to understand and use the SInQUE. |
|  | The initial page where staff are asked to register for the SInQUE. | Staff were asked to enter the organisation they work for in a free-text box. Some found it confusing to know which organisation name they should enter. | Changed response options to a fixed-response, drop-down menu with all housing providers in the borough and 'other' free-text option. | Allows for compilation of service level data and easier to for staff to navigate. |
|  | The page where staff enter details to set up a new SInQUE assessment. | During the introductory paragraph describing the assessment, some staff thought further information on the exact purpose of the questionnaire and how it should be administered would be useful. | Additional guidance on how questionnaire should be administered was added to the introduction paragraph about the SInQUE. | Important contextual information for the questionnaire was explicitly clarified. |
|  | The page where staff enter details to set up a new SInQUE assessment. | Some staff found the wording of the question '*Please select the type of accommodation in which the service user is living from the list below*' to be ambiguous and confusing. | Changed the wording of the question to '*Please select the type of housing support the service user receives from the list below*'. | Clarity of the question was improved. |
| **Changes to the wording of SInQUE questions.** | The section covering 'leisure' questions. | Some staff felt that it was unclear what the '*Other*' option meant in the context of question 3f: '*Over the past year have you been to... Other?*' | Changed the wording of this sub-question to: '*Other leisure activity?*'. | Clarity of the question was improved. |
|  | The section covering 'leisure' questions. | Some staff and service users felt that question 7: '*Do you spend time in pubs or cafés?*' was worded in a way that was potentially inappropriate for people who do not drink alcohol.<sup>a</sup> | Changed the wording of the question to: '*Do you go out for a coffee/drink (e.g. to a café or pub, etc) at least once a week?*' | Clarity and appropriateness of the question was improved. |
|  | The section covering 'social' questions. | Some staff were unsure whether question 9: *'How many people, outside those in your care team, could you confide in?'* related to a professional or personal care team. | Changed the wording of the question to: *'How many people, outside the workers in your care team, could you confide in?'* | Clarity of the question was improved. |
|  | The section covering 'home life/housing' questions. | Some staff thought that question 36: *'What kind of accommodation do you live in?'* was worded ambiguously. | Changed the wording of the question to: *'What type of housing support do you receive?'* | Clarity of the question was improved. |
|  | The section covering 'home life/housing' questions. | A statement alerting users that question 38 had been omitted from the online SInQUE, which read: *'Not relevant for supported accommodation contexts -omitted.'* was confusing. | Changed statement to: *'Question omitted, not included in the online SInQUE.'* | Clarity of the question was improved. |
| **Changes to the SInQUE summary outputs.** | The SInQUE summary report. | Some staff found the spider graph to be confusing to interpret as the numbers summarising scores in each section were not standardised, and therefore it was difficult to tell which domains scored lower than others. | Simplified the spider graph to show percentage/proportion of total score in the graph instead of frequency. | Easier to interpret the graph as sections with different totals became standardised. |
|  | The SInQUE summary report for multiple assessments completed with the same service user. | In the section summarising scores across multiple timepoints, staff thought a visual depiction of this comparison would be useful. | Made comparative bar charts of multiple scores across time with the same service user available on summary report. | Easier to understand and relay the results. |

All changes were made following the initial lab-testing stage, unless indicated otherwise.
<sup>a</sup>Changes were made following field-testing stage.

Overall, changes made to the online SInQUE were relatively few and minor. Following initial lab-testing, some additional information and guidance for users was added, and minor revisions to the wording of questionnaire items were made to improve clarity. Modifications to the visual representation of scores in the summary reports were also made to aid ease of interpretation.

During the field-testing stage, very few suggested changes to improve the usability of the online SInQUE were made by staff or service user participants. Further changes made at this stage included a minor wording adjustment of one question to ensure its cross-cultural appropriateness. We also developed an additional guidance document for managers and commissioners, and an informational leaflet and poster about the SInQUE.

A few participants suggested substantial modifications to the structure and wording of individual items in the SInQUE that were not implemented by the study team. These decisions were made to avoid changes that could disrupt the psychometric properties of the tool, established in the earlier studies, and to preserve its broad scope and logical flow. We also declined to action some suggestions that were outside the remit of the SInQUE tool, such as adding additional or free-text response options to some questions. However, where these suggestions indicated important potential barriers to using the SInQUE, these were noted and integrated into the qualitative analysis and logic model development. A summary of all comments and suggestions that were proposed but not changed after review by the team can be found in the supplementary material (Appendix D).

### Interview thematic analysis

Interviews were analysed using thematic analysis. Primary themes were deductively imposed according to each core element of the ICAMO model – namely intervention, context, actors, mechanisms, and outcomes [38]. Subthemes were inductively analysed within each of these primary themes. The resultant thematic framework is summarised in Table 3. Themes are summarised below with a selection of illustrative quotes. Please see the supplementary materials for further illustrative quotes, for each subtheme (Appendix E).

**Table 3.** Analysis of interviews with users of the online SInQUE: summary of the thematic framework.

| Primary ICAMO themes | Inductive subthemes |
| --- | --- |
| **Intervention:**<br>*Combination of program elements or strategies designed to produce behaviour changes or improve health status among individuals.* | The online SInQUE:<br>- Promotes positive, collaborative discussion<br>- Comprehensive and novel questions<br>- Ability to repeat over time<br>- User friendly design:<br>• Easy to navigate website<br>• Quick to complete<br>• Fixed-response questions<br>• Offers options to choose from<br>• Online format |
| **Context:**<br>*Salient conditions that are likely to enable or constrain the activation of program mechanisms.* | - Relevance of the SInQUE to staff role<br>- Inconsistency in current assessments used across services<br>- Absence of comparably specific assessment<br>- Emergence from the pandemic |
| **Actors:**<br>*The individuals, groups, and institutions who play a role in the implementation and outcomes of an intervention.* | - Staff<br>• Professional knowledge and skills<br>• Professional boundaries<br>- Staff (service user views about staff)<br>• Trusting relationship<br>• Proactivity in offering guidance and support<br>- Service users<br>• Familiarity and comfort with the questions<br>• Individual language/cultural differences of service users<br>- Service users (staff views about service users)<br>• Engagement in the assessment<br>• Existing mental health needs |
| **Mechanisms:**<br>*Any underlying determinants or social behaviours generated in certain contexts.* | Using the online SInQUE can:<br>- Boost service user proactivity and confidence<br>- Identify service users' priorities on social inclusion<br>- Prompt novel, personal conversations<br>- Monitor changes in social inclusion over time<br>- Identify gaps in support available within the organisation and local community |
| **Outcomes:**<br>***Intermediate:** behavioural changes that follow the immediate knowledge change.*<br>***Long-term:** changes such as patient's health status and impact on community and health system.* | - Intermediate:<br>• Improve staff relationship with/understanding of service user<br>• Help to plan more relevant, targeted support for service user<br>- Long-term:<br>• Borough-level improvements/changes in services to support social inclusion<br>• Individual-level benefits for service users' recovery and social inclusion |

### Intervention

In general, staff and service user participants felt that the tool was user-friendly and collaborative. Many noted the ability to repeat the assessment and the online format as being particularly useful, and felt that the website was easy to navigate. The short length of the assessment was also discussed as an important advantage, with both staff and service users commenting that it felt quick to fill out. Participants noted that despite the short assessment length, it still offered a range of interesting, positive, and sometimes unfamiliar questions that felt comprehensive and useful to discuss:

> *“I think that it wasn’t too just baseline, it was a little bit more than that and I think that’s good. Because it gives the option of, ‘Okay, you don’t want something, how can we improve and what is it that you do want that could help you while you’re in our service?’” (212, staff member)*

Interviewees felt that the user-friendliness was aided by providing accessible questions that were straightforward for service users to answer, and that were cross-culturally appropriate for individuals from different ethnic backgrounds. While some participants from both groups felt that the fixed-response options for the questions felt limiting, certain staff members thought that this made the questionnaire more accessible for service users who may otherwise struggle with engagement.

### Context

Staff largely felt that the tool was fitting and relevant to their role in helping support service users, and most did not already use assessments that were highly similar to the SInQUE. Certain staff highlighted a lack of continuity of support workers within their service, and noted that this often made it difficult to build rapport with service users. Some also commented on an inconsistency in assessments used across different services (in the local context where six different provider organisations provided supported accommodation services across the borough). They noted that individual providers currently make their own recommendations on the tools that staff should use:

> *“If it was a standard central assessment that we do in all supported housing, that’s similar, like this for example, it might be beneficial in the long run. But each company has their own policy around it*.*” (205, staff member)*

One staff member noted how the tool felt particularly relevant following the COVID-19 pandemic, as a means to promote engagement among service users after a period of likely sustained social isolation.

### Actors

There were two key actors to consider in the application of the assessment – the staff members who asked the questions and the service users who responded to them.

One staff member felt that the assessment was not particularly relevant in the context of their work in a residential care service, where they had an established relationship with service users and already knew much of the queried information about them. However, this was an outlying view. While most staff members thought the online tool could be suitable and useful for their work, they emphasised the importance of using their professional knowledge and skills to pick when and for whom the assessment would be appropriate. They suggested that service users acutely struggling with their mental health may find it difficult to maintain concentration and engagement with the questionnaire, and others may feel that the assessment is not relevant to them.

Staff also raised the importance of maintaining professional boundaries with service users, and some expressed a concern that certain questions may feel invasive or uncomfortable for service users to answer:

> *“I think there was one quite private like about if they’re in a relationship or something, and that was the only question that made me feel a bit like I’m asking something very personal about a relationship. Because they might not want to say that*.*” (210, staff member)*

Service user participants, however, did not express any similar concern about intrusive questions. They generally indicated that they felt comfortable with the assessment and that they were used to answering personal questions. Both staff and service users highlighted trust between those doing the assessment as a key factor to promote engagement with such questions.

Both respondent groups highlighted the cultural diversity of service users within supported accommodation, and many noted that the tool felt appropriate for those from a variety of religious and ethnic backgrounds. Some service users commented on the importance of the staff member being proactive and taking the time to go through the assessment in detail with them, particularly individuals for whom English was a second language, as further explanation was required for some questions. Various staff suggested that being provided with information about the assessment and its purpose specifically aimed at service users, for instance a guidance leaflet, would be helpful for them to convey the essential information about the assessment.

### Mechanisms

Both participant groups discussed how the tool may boost confidence and proactivity for a wide range of service users, by highlighting specific, achievable ways an individual can improve their social inclusion. They also noted how the assessment encourages service users to open up and enables more profound conversations between them and staff:

*“Yes I found it really interesting, so like because it’s not really topics I would actually talk about. So it gave me a bit of enthusiasm to talk about some of the questions*.*” (409, service user)*

Both groups suggested that it might be particularly useful during key working sessions as a means to get to know an individual better and to identify their support preferences soon after moving into supported accommodation. Participants also noted the value of repeating assessments over time, suggesting that this could be a potentially encouraging way to demonstrate service user progress and to identify gaps in support available. The most frequently suggested time between assessments was 1-3 months, with up to 6 months mentioned as a potential maximum gap.

### Outcomes

Interviewees discussed short and long-term outcomes that they felt the tool could offer. They discussed how the tool enabled targeted and relevant support which prioritised the service user’s interests. Both groups also mentioned the potential for the tool to improve the relationship and understanding between service users and staff:

*“It asks questions where maybe like for your support worker to get a better understanding of you, like even though the immediate thing is highlight areas you can work on, it gives a general overview of how you are*.*” (410, service user)*

Some staff also discussed how prolonged use of the tool could highlight the additional borough-level support that may be needed to improve certain gaps in support, and could also promote service user recovery towards the goal of more independent accommodation.

#### 3. Implementation strategy

Our implementation strategy was developed to encourage use of the SInQUE in the supported accommodation services in the borough and was updated through consultation with clinical staff working in the borough’s community rehabilitation team and supported accommodation service managers. It was further informed by feedback from staff and service user participants during both stages of the study.

Each part of the strategy was developed to target an identified potential barrier to staff using the online SInQUE with service users. We subsequently mapped each component of the strategy to the three broad domains of the COM-B framework of behaviour change [39], to describe whether each element of the strategy was intended to increase staff’s capability, opportunity, or motivation to use the online SInQUE. The complete implementation strategy and the COM-B domain that each component addressed are outlined in Table 4. Strategies related to: enlisting leadership support to encourage supported accommodation staff to use the SInQUE, providing technical guidance and assistance with using the online tool, and developing bespoke summary output reports to reinforce use and increase the organisational benefits of using the SInQUE.

**Table 4.** Summary of implementation strategy to support use of the online SInQUE.

| Activity | Implementation barrier being addressed | COM-B domain |
| --- | --- | --- |
| Study research assistant (SE) and clinical research staff member (MD) visit all supported accommodation services to introduce the SInQUE tool to staff and offer guidance on its use. | Increase awareness of the SInQUE tool among supported accommodation staff and respond to any of their concerns or other problems. | - Motivation<br>- Opportunity<br>- Capability |
| NHS community rehabilitation team manager and clinical researcher (MD) review supported accommodation caseloads to identify suitable service users for a SInQUE assessment and ask their key worker to complete an assessment with them. | Lack of accountability after initially asking all staff to use the SInQUE and staff hesitancy over which service users would be suitable for an assessment. | - Motivation |
| NHS community rehabilitation team manager contacts supported accommodation managers to ask them to support and encourage use of the SInQUE by identified keyworkers within a given timeframe. | Lack of supported accommodation management prioritisation for staff to use the SInQUE. | - Motivation<br>- Opportunity |
| Local Authority commissioners contact all supported accommodation managers to encourage use of the SInQUE within services in the borough. | Lack of supported accommodation management prioritisation for staff to use the SInQUE. | - Motivation |
| Study lead (BLE) and research assistant (SE) attend local Housing Providers Forum meetings to update on the study and encourage use of the SInQUE to all managers and staff present. | Increase visibility and awareness of the SInQUE among supported accommodation managers and respond to any of their concerns or other problems. | - Motivation<br>- Opportunity |
| Study research assistant (SE) offers supported accommodation staff technological support with SInQUE registration and use. | Uncertainty about how to manage the technical process of using the SInQUE. | - Capability |
| Study team develop and circulate a leaflet about the SInQUE for supported accommodation staff to give to service users to help explain the purpose of an assessment. | Uncertainty among some supported accommodation staff about how best to explain the purpose of the SInQUE and engage service users. | - Capability |
| Study team send summary reports to service managers outlining use of the SInQUE and highlighting the areas of social inclusion that are most frequently prioritised and addressed in their service.<sup>a</sup> | Increased awareness among supported accommodation managers of the value offered by the SInQUE for service planning, to encourage them to prompt staff to use it. | - Motivation |
| Study team send summary reports to Local Authority commissioners outlining which services have used the SInQUE the most, and highlighting the areas of social inclusion that are most frequently prioritised and addressed across all services in the borough.<sup>a</sup> | Increased awareness by commissioners of the value offered by the SInQUE for service planning and commissioning, to encourage them to prompt services to use it. | - Motivation |

<sup>a</sup>These actions were planned with service managers and commissioners but not carried out during the 5-month implementation period, due to the small numbers of completed SInQUE assessments.

#### 4. Usage data

In total, 27 staff in the inner London borough registered an account with the online SInQUE. Of the staff who registered, 17 staff members from 6 different supported accommodation providers started or completed a SInQUE assessment with at least one service user. This resulted in 30 completed SInQUE assessments with 28 service users in the borough. This represented just over 10% of the estimated total number of service users living in supported accommodation in the borough. Four of the 28 service users were from residential care, 19 were from 24-hour supported housing, three were from 9 to 5 supported housing, one was from floating outreach services, and one was registered as ‘other’ accommodation type. One staff member from one of the local authority areas where there was no specific implementation strategy registered an account with the SInQUE, however they did not start or complete a SInQUE assessment.

#### 5. Intervention Logic Model

Based on the collective study findings, we developed a logic model to summarise the processes involved in using the SInQUE (Figure 1).

**Figure 1:**
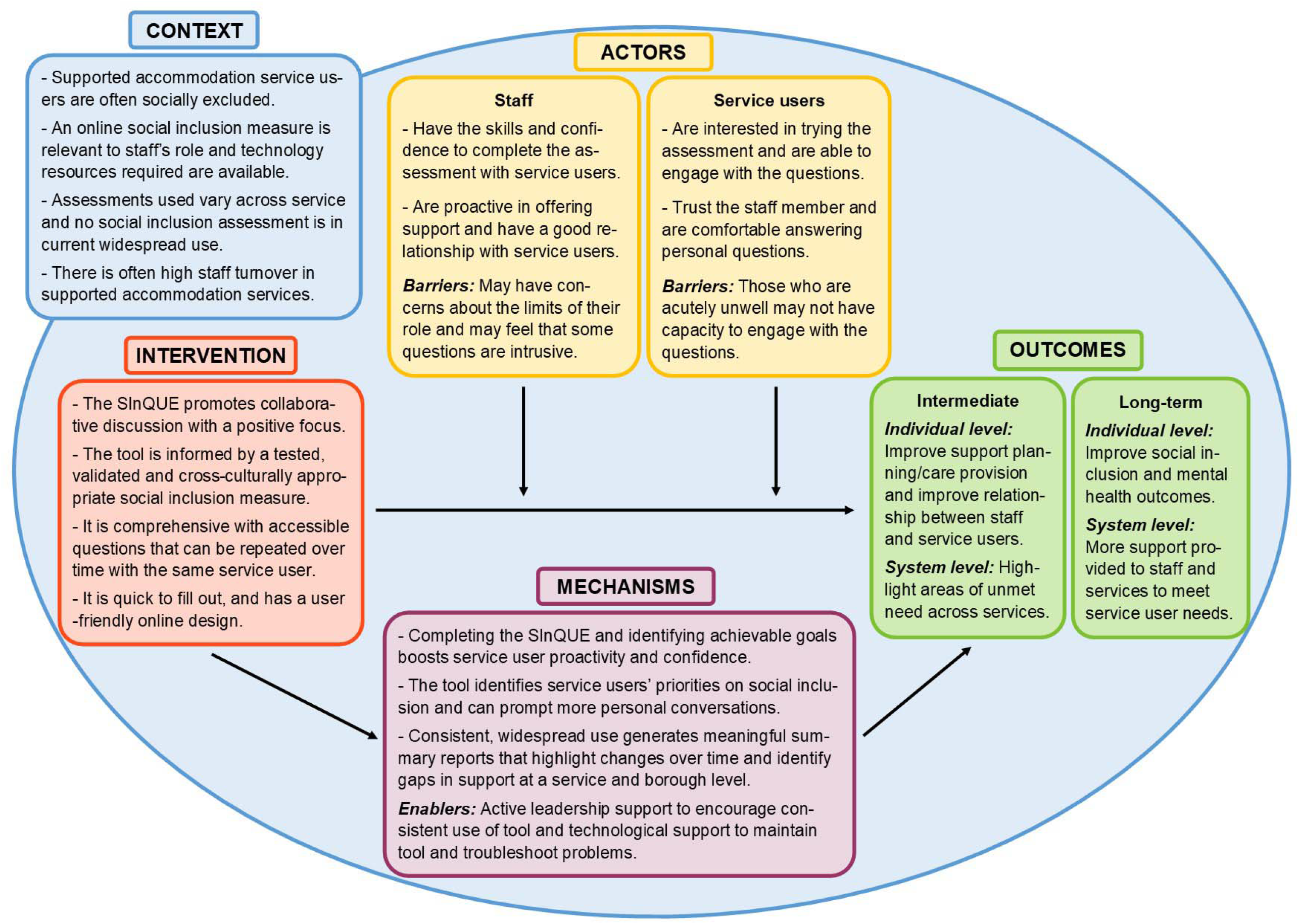
ICAMO logic model for the online SInQUE.

The logic model was informed by the structure of an ‘ICAMO map’ [38]. The model outlines the key aspects of the intervention (I), including its user-friendliness, its comprehensive nature, and that it was based on a validated measure of social inclusion. It also indicates the potential outcomes (O) from using the SInQUE at both an individual and system level, including improved support planning, better relationships, and provision of additional support for staff and services, which in turn may improve social inclusion and mental health outcomes as well as care provision more broadly. These operate within the broader societal context (C) of service users often being socially excluded, that there is high turnover of staff within these services, and a high degree of variation across services in assessment tools that are recommended and in use, rendering the tool useful and relevant to staff’s role.

The key actors (A) in implementing the SInQUE are the staff and service users, who respectively require the skills and proactivity to administer the assessment, and the motivation and trust to engage with the questions. Staff may encounter barriers such as a concern that some questions are too intrusive, and service users struggling more severely with their mental health may lack the concentration or motivation to engage with the questions. The potential outcomes operate through certain mechanisms (M), which include increased service user confidence and the prompting of more in-depth, personal conversations between service users and staff. The tool also identifies more relevant priorities for service users, which may or may not be chosen as an active priority for support by staff due to individual or organisational factors. Persistent and wide-ranging use of the tool could, over time, highlight the aspects of social inclusion that are feasible to work on and those that are regularly not being prioritised.

## Discussion

### Principal results

The online SInQUE was generally perceived as acceptable and potentially useful by supported accommodation staff and service users. This is consistent with findings from previous studies using the SInQUE with other mental health populations [32,34]. Both staff and service users generally found the tool to be user-friendly and relevant, and suggested that it could promote more targeted care planning and improve the relationship between staff and service users. Due to lower uptake of the SInQUE in residential care and floating outreach services, findings relating to the tool’s utility in these settings are less conclusive than for supported housing, where uptake was highest.

Some staff members expressed a concern that certain questions in the SInQUE could be perceived as intrusive for service users, indicating that they did not feel wholly comfortable asking what they perceived to be highly personal questions. This sentiment was not echoed by service users, however, who generally felt that the questions were appropriate and they felt comfortable to answer them. This finding is interesting given that supported accommodation service users have highlighted in previous research the importance of feeling personally understood by staff in their service, and have endorsed a process of familiarisation with staff [25,40].

We found that implementation support is essential in order to promote use of the tool in services, evidenced by the lack of use of the tool in the two regions where the SInQUE was introduced without a concerted implementation strategy. The most effective steps in our implementation strategy were those during which use of the tool was actively endorsed by individuals in leadership positions, particularly service managers and local service leaders. However, even with our concerted implementation strategy, take-up of the SInQUE was only achieved with about 10% of service users living in mental health supported accommodation living in the participating borough, within the 5-month study time period.

### Limitations

We used an established, iterative process of testing and feedback to develop the online SInQUE and determine its real-world acceptability and utility for use in mental health supported accommodation. However, it is important to acknowledge certain limitations of the study.

As aforementioned, uptake of the SInQUE tool was highest in supported housing as compared to residential care homes and floating outreach support. It is unclear whether this discrepancy reflects a greater reluctance from staff and/or service users in residential care and floating outreach support to use the online SInQUE. As proposed by one residential care staff member, it is possible that staff in these services perceived the tool as being less relevant for their role. The discrepancy in partreflects the greater number of 24-hour and 9 to 5 supported housing units in the borough compared to residential care and floating outreach services, with approximately six times as many service users living in supported housing compared to residential care, and nearly twice as many in supported housing compared to floating outreach support. Through the local health service community mental health rehabilitation team, we also had a more direct connection to supported housing teams compared to other service types, which may have further contributed to the imbalance in services where the SInQUE was used.

There were no female service user participants in the qualitative analysis, therefore findings may not be generalisable to women in supported accommodation. It is unclear why it proved more difficult to recruit female participants, though it may reflect the higher proportion of male service users availing of supported accommodation in England -one review suggests that between 68 and 74% of service users are male across all supported accommodation types [41]. Furthermore, as we only tested the SInQUE in one London borough, it is possible that the findings may not be generalisable to other regions.

As the tool was only used with approximately 10% of service users in the borough, the success of our implementation strategy was limited and the low uptake may limit the wider generalisability of our findings. Due to the short time-period and limited scope of the study, it was also not possible to assess whether use of the SInQUE in practice did lead to improved outcomes for service users, or how useful the repeat assessments were over time. As the staff who participated had volunteered to do so, and they chose which service users to complete the SInQUE with, the findings may have been affected by selection bias, and may not accurately reflect all supported accommodation staff and service users’ views.

### Implications for practice

The SInQUE can be recommended as a potentially useful and acceptable tool for use in mental health supported accommodation settings, particularly supported housing services which offer 24-hour or 9 to 5 support, to provide a thorough assessment of social inclusion and support care planning. The tool may help meet an identified wish from service users for more discussion and support with social inclusion and relationships [40]. It was evident during the study that there is currently no universal tool to help with social inclusion in widespread use in mental health supported accommodation, highlighting the potential gap for an assessment tool like the SInQUE. If used widely across supported accommodation services, the online SInQUE has potential to provide benchmarking data and identify service users’ most common priorities for greater social inclusion, to inform service planning and evaluation.

Our findings also suggest that in order for an assessment tool like the SInQUE to be widely used, it is essential to have active leadership endorsement and support for the tool. For example, it may be required for managers and/or commissioners to direct staff to use the SInQUE with service users who are willing, and to reinforce this through team meetings, setting usage targets, or implementing key performance indicators for its use.

### Implications for research

It is important to hear from staff and service users who chose not to use the online SInQUE, to understand their reasons for not using the tool and highlight barriers for using the tool that we may have missed in the present study. It would be useful to conduct further testing of the tool in residential care and floating support supported accommodation settings, to better determine the utility of the SInQUE in these service types. It would also be useful to examine the utility of the SInQUE in other population groups within different service types, to determine whether the tool may be useful in additional settings.

Future research is necessary to establish the level of uptake of the SInQUE that can be achieved in supported accommodation over a longer time period, and to potentially establish more effective means of implementation support. A longer-term study is also needed to establish whether the possible benefits from using the SInQUE that were mentioned by staff and service users are achievable through use of the tool, and how any potential outcomes may vary over time. A hybrid implementation-evaluation study would address these queries, to determine the effectiveness of the SInQUE tool as an intervention for social inclusion, and to establish a precise implementation strategy for widespread uptake of the tool in supported accommodation. Further research using the SInQUE is also warranted to examine service user needs relating to social inclusion and to identify any additional barriers to addressing these needs in supported accommodation services. Such research could be used to inform the development of a future complex intervention to support social inclusion in supported accommodation services.

## Data Availability

All data produced in the present study are available upon reasonable request to the authors.

## Acknowledgements

This paper presents independent research funded by the National Institute for Health and Care Research, School for Social Care Research. The views expressed are those of the authors and not necessarily those of the National Institute for Health and Care Research or the Department of Health and Social Care.

We would like to thank the supported accommodation managers, staff, and service users who supported and participated in this study. We would also like to thank members of our expert advisory group and the UCL Service User Research Forum (SURF) for their advice and feedback throughout the study.

## Conflicts of Interest

None declared.

## Abbreviations

SInQUE: Social Inclusion Questionnaire User Experience
QuIRC-SA: Quality Indicator for Rehabilitative Care - Supported Accommodation
NHS: National Health Service
TIDieR: Template for Intervention Description and Replication
UCL: University College London
ICAMO: Intervention-Context-Actor-Mechamism-Outcome
REC: Research Ethics Committee
SURF: Service User Research Forum

## Supplementary Files

Appendix A: Full description of the online SInQUE using the TIDieR checklist for reporting interventions

Appendix B: Lab-testing topic guide

Appendix C: Field-testing topic guides (for staff and service users)

Appendix D: Explanation for suggested changes during lab and field-testing that were not made after team review

Appendix E: Interviews with staff and service users: Illustrative quotes for each inductively derived subtheme

## Notes

### Competing Interest Statement

The authors have declared no competing interest.

### Funding Statement

This study was funded by the National Institute for Health and Care Research, School for Social Care Research.

### Author Declarations

Research Ethics Committees of University College London (reference: 6711/002) and London Camden and Kings Cross National Health Service (reference: 21/LO/0657) gave ethical approval for this work.

