## Appendix A for "Development and preliminary testing of an online tool to assess social inclusion and support care planning in mental health supported accommodation"

Appendix A – Full description of the online SInQUE using the TIDieR checklist for reporting interventions

| ***TIDieR checklist item*** | ***SInQUE details*** |
| --- | --- |
| **1. Brief name**  *Provide the name or a phrase that describes the intervention*. | The Social Inclusion Questionnaire User Experience (SInQUE: <https://sinque.org.uk/>). |
| **2. Why**  *Describe any rationale, theory, or goal of the elements essential to the intervention.* | The SInQUE is an online tool to assess social inclusion and support care planning for people with severe mental illness living in supported accommodation. It is designed to be delivered by supported accommodation staff as part of routine care and, to be completed collaboratively with service users. |
| **3. What: materials**  *Describe any physical or informational materials used in the intervention, including those provided to participants or used in intervention delivery or in training of intervention providers.*  *Provide information on where the materials can be accessed (e.g. online appendix, URL).* | Guidance materials for managers, staff, and service users are provided on the online SInQUE website (<https://sinque.org.uk/>). Information includes instructions at the start of the assessment, a guidance document for service managers and commissioners, an informational leaflet for staff, and a poster with information for service users. There is also a frequently asked questions section and background information about the development of the SInQUE on the website. |
| **4. What: procedures**  *Describe each of the procedures, activities, and/or processes used in the intervention, including any enabling or support activities.* | Supported accommodation staff are required first to register an account with the online SInQUE using their work email address and a password. Once they have registered, they can then start a SInQUE assessment with one of their clients. No personal information of the client is entered or stored by the website – by starting a new assessment a random ID number is generated and only the staff member doing the assessment will know which number belongs to each client. The assessment consists of 46 questions taken from the psychometrically validated SInQUE questionnaire [32], which are answerable using fixed response options in a drop-down menu. The questions correspond to nine different areas of social inclusion: leisure activities, social life, home life/housing, transport, health, religious/cultural activities, civic duties, crime victimisation, and education/employment. The assessment can be saved and returned to later if an individual wants to take a break during the questions. When the questions have been completed, the staff and client are then given an overall score of social inclusion and a list of areas that the client said they would like more support with their social inclusion, and they are prompted to choose up to three of these to work on together as part of their care planning. A report is also produced, summarising their responses. Staff can repeat the assessment with service users in the future, to see whether their social inclusion has changed. |
| **5. Who provided**  *For each category of intervention provider (e.g. psychologist, nursing assistant), describe their expertise, background and any specific training given.* | A SInQUE assessment can be delivered by any staff member working in mental health supported accommodation. No specific training was required for them to use the SInQUE. |
| **6. How**  *Describe the modes of delivery (e.g. face-to-face or by some other mechanism, such as internet or telephone) of the intervention and whether it was provided individually or in a group.* | The intervention was delivered via an online website by the staff member, while present face-to-face with one service user. The staff member read out the questions like an individual interview and input the service user’s responses into the website. |
| **7. Where**  *Describe the type(s) of location(s) where the intervention occurred, including any necessary infrastructure or relevant features.* | The intervention was delivered in supported accommodation services, typically in a private room where there was access to a computer. The intervention was also delivered in some service user’s homes, depending on where the staff member usually met with them. It can be delivered in any location that is suitable for the service user, requiring only a computer or mobile device and an internet connection. |
| **8. When and how much**  *Describe the number of times the intervention was delivered and over what period of time including the number of sessions, their schedule, and their duration, intensity or dose.* | The intervention was delivered 30 times throughout the study, with 28 different service users. It was delivered across a 5-month period. Due to the time constraints of the study, the intervention was typically delivered in one session by the staff member, with a recommended time to wait before repeating of approximately 3-6 months. Each assessment typically takes around 15/20 minutes. |
| **9. Tailoring**  *If the intervention was planned to be personalised, titrated or adapted, then describe what, why, when, and how.* | N/A. |
| **10. Modifications**  *If the intervention was modified during the course of the study, describe the changes (what, why, when, and how).* | Minor adjustments were made to the SInQUE following two rounds of testing with staff and service users. Modifications included changing the wording of a small number of items in the questionnaire to improve clarity, adding additional information and guidance for staff using the questionnaire, and changes to the visual representation of scores in the summary reports to aid ease of interpretation. |
| **11. How well: planned**  *If intervention adherence or fidelity was assessed, describe how and by whom, and if any strategies were used to maintain or improve fidelity, describe them.* | N/A. |
| **12. How well: actual**  *If intervention adherence or fidelity was assessed, describe the extent to which the intervention was delivered as planned.* | N/A. |
