## Appendix B for "Development and preliminary testing of an online tool to assess social inclusion and support care planning in mental health supported accommodation"

Appendix B – Lab-testing topic guide.

**Topic guide for “Think Aloud” exercise and interview with staff participant**

**Version 1, 12/04/21**

**A. Information about you**

[Researchers: record and store this information with the participant’s consent form: keep separate from the data collected in sections B and C]

- Gender (Male; Female; Trans/other; prefer not to say)
- Age (18-30; 31-50; 51+; prefer not to say)
- Ethnicity (White/White British; Black/Black British; Asian/Asian British; Mixed ethnic groups; Other ethnic groups; prefer not to say)
- Housing organisation you work for
- Type of supported housing you usually work in

1) Residential care (24-hour staffed; communal facilities; resident does not have a tenancy; stay is not time-limited; high support)
2) Supported housing - 24 hour support (self-contained, time-limited tenancy within a shared house; staff present 24 hours; medium support)
3) Supported housing – not 24 hour support (self-contained, time-limited tenancy within a shared house; staff based on site but not all 24 hours; medium support)
4) Floating outreach services (visiting support from housing service staff to people living in self-contained, permanent individual tenancies; lower support)

- How long have you worked in supported housing services? (less than 2 years; 2-5 years; 6-10 years; more than 10 years)

**B. Think Aloud exercise**

*We are now going to ask you to use the online SInQUE assessment tool. First you will have to register as a staff member to use the tool. Then, you will have to complete a new profile for an imaginary service user, and complete an assessment, as if you were doing it with them. Finally, we will ask you to access the feedback from the assessment, which will include some space for you to record agreed priorities for future work.*

*At all stages, we would like you to “think aloud” and share your thoughts about how you are finding it navigating the online tool, and what you think about its relevance or potential usefulness for your work. The purpose of this exercise is not to get suggestions about new or different questions for the SInQUE assessment, which has been developed and tested with service users in previous research. We want to get your feedback about using the online tool, from your perspective ass a staff member in a supported accommodation service. This is a new online tool: it may not be as clear and user-friendly as it needs to be. That’s what we want to find out: we are testing the online tool, not you, so please do let us know any points where you are not sure what to do or why you are being asked to do something.*

*We will ask you to complete the online assessment for an imaginary resident: please do not provide information about any specific, real person.*

**i) User registration:** *If you’re ready, please can we ask you to access the web page for the online SINQUE. As you are doing this,*

- Tell us how easy or difficult you are finding it to register
- Tell us about anything you are asked which surprises or worries you

**ii) New resident registration:** *Now we’d like you to set up an account for a new resident who hasn’t had a previous online SInQUE assessment. As before:*

- Tell us how easy or difficult you are finding it
- Tell us about anything you are asked which surprises or worries you

**iii) Online assessment completion:** *Now we’d like you to complete the online SInQUE social inclusion assessment, as if you were completing it with the resident you’ve just registered. As you are doing this:*

- Tell us how easy or difficult you are finding it
- Tell us about anything you are asked which surprises or worries you, or you think might worry the resident you were completing the assessment with

**iv) Getting the report from the assessment:** *Now we’d like you to access the report generated from the assessment you have completed. Please ask to see for all the information you are offered. As you are doing this:*

- *Tell us how easy or difficult you are finding it to access the report*
- *Tell us what you think about the information given to you in the report*
- *Tell us about anything you were hoping to see which isn’t provided here*

**C. Post-completion interview**

**1. How did you find using this online assessment?**

Prompts:

- Was it easy to work out what to do; were the steps easily explained?
- How burdensome did you find it?

**2. Would you like to use this online assessment with residents in your actual work?**

Prompts:

If NO, explore barriers to using it (probe available time, relevance to work role, perceived acceptability to resident)

If YES, explore perceived usefulness (probe best timing; with all or only some residents; how the report information could be used)

**3. What, if anything, could make this online assessment tool more useful for you or other staff working in mental health supported accommodation services?**

Prompt:

- Any improvements to the instructions or guidance to make it easier to navigate?
- Any changes to the language or tone which would increase acceptability?
- Any unnecessary or off-putting questions or stages to the user process?
- Any extra information which could improve the usefulness of the feedback report?

**4. Please tell us any other reflections about this online social inclusion tool and how it could be used in supported accommodation services.**
