## Appendix C for "Development and preliminary testing of an online tool to assess social inclusion and support care planning in mental health supported accommodation"

Appendix C – Field-testing topic guides (one for staff and one for service users).

**Topic Guide for staff interviews
Version 1, 31/07/2021**

*Thank you for taking part in this interview. I’ll ask you about your experience of doing the online SInQUE assessment, and any ways you think it could be improved. I’ll start recording now. If you want to take a break at any point during the interview, or to stop, just let me know.*

**1. Please can you tell me how many service users you have done a SInQUE online social inclusion assessment with?**

**2. How did you find using the social inclusion assessment?**

Prompts:

- Were you and the service users clear what it was for and why you were doing it
- Duration and burden of completing all the questions (did people complete it in one go? Was there anything off-putting or difficult?)
- Pros and cons of an online tool
- Choosing areas where you would like help to be more socially included (did you and the service user agree on priorities to work on together?)
- Looking at the feedback report from the assessment

**3. What ways, if any, could the online social inclusion assessment be improved?**

Prompts:

- Changes to the information provided about the assessment
- Changes to the feedback report

**4. What, if anything, was a barrier to you using the online tool or made you reluctant to do so?**

Prompts:

- Availability of technology
- Concerns about using an external online tool
- Available time
- Work role and priorities
- Service users’ attitudes and enthusiasm

**5. What impacts, if any, were there from doing the online social inclusion assessment with service users?**

Prompts:

- Any impact on their relationship with you and the sorts of conversations you have?
- Any change to care planning or the sorts of help you offer?

**6. Do you think this online assessment could be useful for other people living in supported accommodation services?**

Prompts:

- When is the best time to do an assessment like this?
- Any groups of people for whom or circumstances in which the assessment would be particularly helpful, or unhelpful
- Cultural appropriateness for different groups?

**7. Is there anything else you would like to tell us about your views of the online social inclusion assessment and your experience of using it?**

**Topic Guide for service user interviews
Version 1, 31/07/2021**

*Thank you for taking part in this interview. I’ll ask you about your experience of doing the online SInQUE questionnaire, and any ways you think it could be improved. I’ll start recording now. If you want to take a break at any point during the interview, or to stop, just let me know.*

**1. What was it like doing the social inclusion questionnaire with your supported accommodation worker?**

Prompts:

- Were you clear what it was for and why you were doing it?
- Duration and burden of completing all the questions (did you complete it in one go?)
- Was there anything off-putting or difficult?
- Pros and cons of an online questionnaire
- Considering areas where you would like help to be more socially included
- Looking at the feedback report from the questionnaire

**2. What ways, if any, could the online social inclusion questionnaire be improved?**

Prompts:

- Usability of the online tool
- The information provided about the questionnaire
- Changes to the feedback report

**3. What impacts, if any, do you think you got from doing the online social inclusion questionnaire with supported accommodation staff?**

Prompts:

- Any impact on their relationship with you and the sorts of conversations you have?
- Any change to care planning or the sorts of help you are receiving?

**4. Do you think this online questionnaire could be useful for other people living in supported accommodation services like you?**

Prompts:

- When is the best time to do an online questionnaire like this?
- Any groups of people for whom or circumstances in which the questionnaire would be particularly helpful, or unhelpful

**5. Is there anything else you would like to tell us about your views of the online social inclusion questionnaire and your experience of using it?**
