## Appendix D for "Development and preliminary testing of an online tool to assess social inclusion and support care planning in mental health supported accommodation"

Appendix D - Explanation for suggested changes during lab and field-testing that were not made after team review.

| ***Location of issue*** | ***Suggested change/explanation of problem*** | ***No. of participants with issue*** | ***Justification for not changing*** |
| --- | --- | --- | --- |
| Leisure questions - *Q3: Over the past year have you been to… Cinema? Theatre? Concerts? Galleries/exhibitions? Sporting matches/events? Other?* | Unclear if one can select multiple options or should just pick one from the list. | 2 | Participants reached correct conclusion through trial and error. |
| Leisure questions page - *Q4: Have you taken a holiday in the past year?* | Unsure whether COVID should be taken into account for this question. Also unsure whether this included breaks like Christmas, for example. | 1 | This can be subjective - want questions to be self-defined and inclusive. |
| Social questions - *Q8: Do you have anyone you would call a close friend?* | Unclear what constitutes a close friend and whether this would include family members as well. | 2 | This can be subjective - want questions to be self-defined and inclusive. |
| Social questions - *Q10: Are you on friendly terms with any of your neighbours?* | Unclear whether this refers to neighbours before the resident came to the service, or neighbours since they have moved in. | 2 | This can be subjective - want questions to be self-defined and inclusive. |
| Education/economic/employment - *Q23: Do you feel poor?* | Proposed that word 'poor' has negative connotations. Fear that it could be perceived as insulting. | 7 | No service users expressed concerns about the appropriateness of this question. |
| Education/economic/employment – *Q26: Do you have any savings or investments?* | Proposed clarifying a minimum amount to quantify what counted as ‘savings’. | 1 | This can be subjective - want questions to be self-defined and inclusive. |
| Transport - *Q28: Do you own a car or motorbike/scooter?* | Suggested that it would be useful to find out if the client has (currently) or had (in the past) a licence, suggested adding a pop-up box. | 1 | Not within the goal of the questionnaire. |
| Health - *Q30: Do you have any serious long-term physical health problems that limit what you can do in any way?* | Proposed offering examples of what would constitute long-term health conditions. | 1 | This can be subjective - want questions to be self-defined and inclusive. |
| Crime victimisation - *Q33: Have you or do you feel you have been a victim of crime or anti-social behaviour/harassment within the past year?* | Proposed offering examples of what would constitute antisocial behaviour. | 1 | This can be subjective - want questions to be self-defined and inclusive. |
| Home life/housing page - *Q42: Would you be able to replace old or worn-out clothes when you want to?* | Unsure whether this question is worded appropriately and/or whether it is relevant. | 3 | No service users expressed concerns about the appropriateness of this question. |
| SInQUE Assessment Results - *Q: Please select up to three of these areas (if available) to work on together to increase this person's social inclusion* | Suggested including an additional 'other' option to input a goal defined by the service user themselves, if goal was not mentioned on list. | 2 | Do not want answers that are not focused on social inclusion. |
| Report page - *Domain score table.* | Unclear how the numbers should be interpreted and what constitutes a good or a bad score. | 6 | This can be subjective - want this interpretation to be self-defined and inclusive. Information on this is provided on the SInQUE site. |
| General. | Proposed adding a ‘sometimes’ or free-text response option to a range of yes/no questions. | 3 | This would interfere with the psychometric properties of the measure. |
| General | Proposed having an adaptable version of the questionnaire, with the option to cut out certain questions depending on the service user. | 3 | This would interfere with the psychometric properties of the measure. |
| General. | Suggested having a more direct way to identify which assessment belonged to each service user, with optional identifying information. | 2 | Due to data protection and privacy, no potentially identifiable service user information should be input into the SInQUE. |
| General. | Suggested option to have paper copy for participants who do not want to use a computer website. | 3 | Not within the goal of this questionnaire, which aims to be fully online. |
| General. | Suggested having an option for the service user to complete the questionnaire by themselves. | 1 | Not within the goal of this questionnaire, which is designed to be completed collaboratively between service user and staff. |
