## Appendix E for "Development and preliminary testing of an online tool to assess social inclusion and support care planning in mental health supported accommodation"

Appendix E – Interviews with staff and service users: Illustrative quotes for each inductively derived subtheme.

| *Primary ICAMO themes* | *Inductive subthemes* | *Explanation of subtheme* | *Illustrative quotes* |
| --- | --- | --- | --- |
| Intervention | Promotes positive, collaborative discussion | The tool felt more conversational than other assessments, with a positive, solution-based focus. | *“Yes I think that sitting down and having it for both myself and the client was useful as they could see it as well as having me read it out to them.” (201, staff member)*  *“I just generally feel like it was a positive outlook. Because it was more like, ‘what can we do to help?’ not just asking, ‘what is it that you like?’” (212, staff member)* |
|  | Comprehensive and novel questions | It covered all the important aspects implicated in social inclusion, including some questions that both groups had not considered previously. | *“I think the questions covered everything that could be a problem in this sort of situation [pause] I mean I can’t think of any areas it was lacking, so.” (407, service user)*  *“I think maybe the assessment had a little bit more in certain topics. But I can’t remember exactly what it was but I do remember seeing some and thinking ‘oh, actually we haven't asked them that’”. (212, staff member)* |
|  | Ability to repeat over time | Built-in aim to repeat the assessment was useful for encouraging progress and identifying changes. | *“In this guy’s case I would definitely want to do another one because all those three variables have changed and there has been progress that’s been made.” (205, staff)* |
|  | User-friendly design | The website was easy to navigate, and interviewees liked the online format. The questions were also accessible and quick to fill out, aided by the provision of closed options. | *“Well, it was very easy, I wasn’t asked very many questions, it only lasted about 10 minutes, 15 minutes something like that.” (404, service user)*  *“The advantages are that it’s accessible, it’s online so different services can use it. It’s online so they have got that access more easily than like paper, it’s not lost.” (204, staff)* |
| Context | Relevance to staff role | Staff felt the tool was fitting with their role and the goals they want to work on with their clients. | *“It’s not just the housing that people have got to deal with, the reason why they come to support it’s because they’ve got other issues that’s preventing them from being able to deal with things themselves. So, something like this may open their eyes to what can make certain situations better.” (211, staff)* |
|  | Inconsistency across services | Staff noted that there was no standardised assessment used across services, and that each had their own policy on what to use. | *“If it was a standard central assessment that we do in all supported housing, that’s similar, like this for example, it might be beneficial in the long run. But each company has their own policy around it.” (203, staff)* |
|  | Absence of comparably specific assessment | No staff identified assessments in use that were comparably focused on social inclusion or were similarly designed. | *“We don’t have like questionnaires we do, no. Not this kind of assessment. […] There’s nothing like that we do this way like they borrow the computer or laptop or whatever and we type in with them.” (210, staff)* |
|  | Language/cultural differences of service users | Both groups generally found the assessment to be culturally neutral and acceptable across different ethnic groups. | *“Nothing came up when I was doing it as well where I thought it wouldn’t fit in with certain groups like religious or racial whatever. It was all pretty neutral really. There wasn’t really anything specific to what would even implicitly seem specific to a certain group which was good.” (208, staff)* |
|  | Emergence from the pandemic | Staff identified the utility of the tool as a means to encourage societal reintegration after a period of particular isolation during the pandemic. | *“But we need more services like this [SInQUE] in order to help people out of it because they are not going to do it themselves. They have been stagnating for two years, here’s how to get the ball rolling again.” (205, staff member)* |
| Actors | Staff:   - Professional skills and knowledge - Professional boundaries | Staff identified the importance of having the skills to complete the questionnaire with service users, in trusting their professional knowledge on whom to complete it with, and the importance of maintaining their professional boundaries with service users. | *“It takes a whole lot of skills for you to be able […] to do a questionnaire with an individual that can only stay focused for five minutes.” (206, staff)*  *“I think there was one quite private like about if they’re in a relationship or something and that was the only question that made me feel a bit like, I’m asking something very personal about a relationship. Because they might not want to say that.” (210, staff)* |
|  | Staff (service user views):   - Trusting relationship - Proactivity in offering support | Service users noted the importance of being of having a good relationship with the staff who does the assessment and being able to trust them. They also noted the staff’s responsibility to be willing and proactive in offering support to them. | *“It also helps because it’s with my worker, my support worker [Staff name]. So if it’d been a random person and you’ve never met them maybe it would’ve been a bit more cold so it wouldn’t have been as useful.” (410, service user)*  *“It all depends on the person who’s offering the support and how good they are, I know it sounds bad. I mean you can have all the forms and the surveys in the world but it’s what does that support worker do with them.” (410, service user)* |
|  | Service users:   - Familiarity and comfort with the questions - Individual preferences | Service users felt that the questions were generally familiar and they were comfortable answering them. They also noted that people’s willingness to engage with the questionnaire was likely to vary due to individual differences and preferences. | *“I think they're just personal questions and I didn’t really have a problem with it, I don't think I got like- yes, I wouldn't regret it. I don't keep secrets or anything in here, I don't have anything to hide really, so.” (402, service user)*  *[On whether the questionnaire would be useful for other people] “It depends, it depends on how the person feels because sometimes we have ups and downs.” (411, service user)* |
|  | Service users (staff views)   - Engagement with the assessment - Existing mental health needs | Staff discussed how some service users may struggle to engage with the questionnaire due to interest, capacity, and motivation. Some also noted that the questions may not be as relevant for service users with more prevalent or acute mental health problems. | *“I think some of them might be willing to do it but not able to concentrate and think about something, focusing on something for a long time- although it’s not long, but still we are talking about people that mentally have difficulties, so I think it’s a bit of a mix of like motivation, like actually capability.” (210, staff member)*  *“Because I think a lot of these things are - because this is residential care, so I think a lot of these things are for people who are much further along in their recovery than people we’ve got here.” (207, staff member)* |
| Mechanisms | Boost service user proactivity and confidence | Helped identify achievable, manageable goals for improving social inclusion, many of which the service users could act on themselves. | *“This sort of opens up to them what they can do to help themselves and start moving forward and that releases us a bit to give them more leeway for themselves. So, like, ‘Oh, can you make that phone call?’ When they see that they can do things, I think that’s going to be the benefit for this online survey.” (211, staff member)* |
|  | Identify service users’ priorities on social inclusion | Highlighted specific things that service users wanted support with and things that they were not interested in. | *“Yes, that makes sense because you want to know what you need help on so, they can show you the points.” (401, service user)* |
|  | Prompt novel, personal conversations | Enabled conversations on personal topics that staff and service users had not discussed before. | *“Yes I found it really interesting, so like because it’s not really topics I would actually talk about. So it gave me a bit of enthusiasm to talk about some of the questions.” (409, service user)* |
|  | Monitor changes in social inclusion over time | Useful to consider how a service user’s social inclusion does or does not change over time, and if not identify any issues that are causing problems. | *“I like that bit, the bit where you’ve done more than one and it can show you your score over time. Now that is useful because you can see whether it’s going up or down. If it goes down than maybe it leads the support worker to think ‘what happened in that period.’” (410, service user)* |
|  | Identify gaps in support available | Helpful to highlight more long term where gaps in resources for social inclusion may be. | *“I think it would be useful when […] you are kind of looking at areas that are not doing so great how do we maximise some of those potentials? What resources do we need to improve within the borough to allow those things to happen?” (203, staff member)* |
| Outcomes | Improve relationship with/understanding of client | Staff can gain a better understanding of the service user and their needs, and thus improve this relationship. | *“It asks questions where maybe like for your support worker to get a better understanding of you, like even though the immediate thing is highlight areas you can work on, it gives a general overview of how you are.” (410, service user)* |
|  | More relevant, targeted support for client | Service users can get involved in particular services or activities that they identified as having an interest in. | *“We sat down after the assessment and said, ‘Okay, what days would you like to do this? How would you like to do this, where would you like to go?’, etc. And we’re actually putting that in place for her because it was to do with going to the café which she likes doing.” (212, staff member)* |
|  | Borough level improvements/changes | Borough-level gaps in support can be identified and can then be targeted to improve support at a wider level. | *“Its local government isn’t it and voluntary services to put things in place for better social inclusion. […] It’s about, you know, it’s all well and good having a good spider web, but how do we improve it when the resources are not there?” (203, staff member)* |
|  | Help with recovery | Service users can improve their social inclusion and recovery, to potentially move to lower levels of supported accommodation. | *“It’s like the whole point of supported accommodation is ideally to move one person from here to […] a less supported place basically. So if we can, if the SInQUE basically can help facilitate that move then yes absolutely because that’s the whole point of our job here really.” (208, staff member)* |
